# A randomised-controlled Phase I de-escalation trial of Molnupiravir and Nirmatrelvir/Ritonavir combination for mild-moderate SARS-CoV-2 infection

**DOI:** 10.1101/2025.05.01.25326797

**Authors:** Saye H. Khoo, Richard FitzGerald, Christopher J. Edwards, Shazaad Ahmad, Geoffrey Saunders, Laura J. Else, Victoria Shaw, Pavel Mozgunov, Josh Northey, Laura Dickinson, Emma Knox, Amanda Buadi, Colin Hale, Helen E. Reynolds, Calley Middleton, Katie Bullock, Lauren Walker, Michelle Tetlow, Rebecca Lyon, Jennifer Gibney, Alieu Amara, William Greenhalf, Abigail Burdon, Jan Dixon, Thomas Jaki, Justin Chiong, David G. Lalloo, Andew Owen, Michael Jacobs, Thomas Fletcher, Gareth Griffiths, the AGILE CST-8 team

## Abstract

**Background:** The AGILE CST-8 (NCT04746183) Phase I de-escalation trial evaluated the safety and tolerability of combination molnupiravir and nirmatrelvir/ritonavir for mild-moderate COVID-19.

**Methods:** Adult out-patients with SARS-CoV-2 infection within five days of symptoms were randomly assigned 2:1 to receive molnupiravir (starting at 800mg twice daily (BD) reducing to 600mg and 400mg if necessary) in combination with nirmatrelvir (300mg)/ritonavir (100mg) BD for 5 days versus Standard of care. Using a dose de-escalation, open-label, Bayesian adaptive Phase I trial a combination dose was considered unsafe if the probability of 30% or greater dose-limiting toxicity risk (DLT - the primary outcome) over standard of care was 25% or higher. Secondary endpoints included tolerability, clinical progression, pharmacokinetics and virological responses.

**Findings:** Of 49 participants screened, 24 were enrolled (16 combination, 8 standard of care) between January 2023 and September 2023. For the primary endpoint, to day 11, no participant starting molnupiravir at 800mg (BD) in combination with nirmatrelvir/ritonavir reported a DLT by day 11 (primary endpoint) or by day 29; dose de-escalation was not required. No participants reported severe adverse events (grade>=3). Although proportion of swab PCR negativity at day 5 and day 11 were not statistically different, faster initial viral clearance was observed with treatment. Penetration of nirmatrelvir into saliva, nasal secretions and tears was 19%, 65% and 91% that of plasma.

**Interpretation:** Molnupiravir in combination with nirmatrelvir/ritonavir was safe and well-tolerated; later phase trials should evaluate combination therapy at currently recommended doses for each drug.

**Research in Context:** *Evidence before this study:* SARS-CoV-2 antivirals such as nirmatrelvir/ritonavir, remdesivir and molnupiravir have been associated with clinical benefit and faster viral clearance when given early to patients at high-risk of severe disease. However, evidence has accrued to suggest that virus persists in tissues (e.g. gastrointestinal tract) and in blood even after clearance from the upper respiratory tract. Moreover, persistent viral infection is recognised in individuals who are the most severely immunosuppressed, (e.g. transplant recipients, those with haematological malignancy, or receiving B-cell depleting therapies) suggesting greater antiviral potency is needed. A PubMed search (26^th^ April 2025) using the terms ‘SARS-CoV-2’ AND ‘antiviral’ AND ‘combination therapy’ revealed descriptions of retrospective and prospective of case series of combination antiviral therapy, usually given to severely immunocompromised patients. Once prospective study from Naples evaluated the monoclonal antibody sotrovimab in combination with antiviral small molecules, without any controls. There were no randomised trials of combination antiviral therapy for SARS-CoV-2, and little recognition that use of agents at their licensed doses might not be be equally safe when combined.

*Added value of this study:* Coadministration of molnupiravir and nirmatrelvir/ritonavir (at fully licensed doses) is safe and well-tolerated. Although numbers were small and no differences were detectable in time to achieving PCR-negative swabs, we observed a significantly faster rate of initial viral clearance with combination therapy (compared with no antiviral therapy) using a biexponential model to examine the initial ‘fast’ decay in viral elimination.

*Implications of all the available evidence:* Molnupiravir plus nirmatrelvir/ritonavir represents an oral-based combination regimen for SARS-CoV-2 infection which should be evaluated against conventional monotherapy, particularly in patients with severe immunosuppression. The use of an adaptive Bayesian dose de-escalation design offers advantages in statistical precision and efficient decision-making for combination antiviral therapy.

## INTRODUCTION

Antiviral treatment for SARS-CoV-2 infection is focussed on high-risk individuals in the UK.^1^ In profoundly immunosuppressed patients, chronic persisting or relapsing SARS-CoV-2 infection despite currently approved antiviral treatments (e.g. nirmatrelvir/ritonavir, molnupiravir or remdesivir) may create a window of opportunity for the evolution of new variants, and the emergence of drug-resistant mutations where antiviral selective pressure is applied. Indeed, such a scenario has been described for remdesivir and nirmatrelvir^2–6^ although the prevalence of drug resistance is not currently widespread.^7^

The failure of current antiviral therapy to reliably eradicate chronic infection in these patients, coupled with evidence for a more prolonged persistence of SARS-CoV-2 in blood^8^ and gut tissues suggests that greater antiviral potency and a higher genetic barrier to resistance may be required. Combinations with two^9^ or three^10^ antivirals have been empirically administered for persistent SARS-CoV-2 infections, but combination therapy has not been systematically evaluated in randomised studies.

Combination therapy with molnupiravir and nirmatrelvir/ritonavir is associated with greater *in-vitro* potency^11^ and greater reductions in viral shedding and replication and reduced lung pathology in animal models.^12,13^ However, such an approach is not without risk: any increase in toxicity cannot be adequately characterised in such complex patients, the efficacy of such combinations needs to be properly tested, and the correct use of each drug at full dose within any combination cannot be automatically assumed. Further, in the case of combinations involving molnupiravir (which induces hypermutation) there is at least a theoretical risk of inducing mutations which become enriched for drug resistance under selective pressure from a second antiviral drug. In macaques treated with molnupiravir, mutations in 3CL protease (including T21L, L50F, A173V, and P252L; conferring resistance to nirmatrelvir) were observed but combination molnupiravir plus nirmatrelvir was not associated with selective enrichment of these mutations.^13,14^

AGILE is the UK early-phase trials platform evaluating experimental COVID-19 antivirals^15^ through a master protocol and individual candidate-specific trials. AGILE undertakes dose optimisation using a Bayesian model to characterise the relationship between dose and toxicity. In this study (CST-8), we undertook an open-label, randomised, Phase I dose de-escalation trial to assess the safety and tolerability of drug combination of molnupiravir and nirmatrelvir/ritonavir in patients with mild-moderate SARS-CoV-2 infection.

## METHODS

### Trial design and Oversight

The AGILE CST8 trial (<u>NCT04746183</u>) was an open-label, randomized, controlled, de-escalation Bayesian adaptive Phase I trial in adult out-patients with COVID-19 within 5 days of symptom onset with the primary objective to determine the safety and tolerability and recommended phase II dose of molnupiravir in combination with nirmatrelvir/ritonavir. Eligible participants were men and women aged ≥18Lyears with lateral flow positive SARS-CoV-2 infection who were within fiveLdays of symptom onset, free of uncontrolled chronic conditions, ambulant in the community with mild or moderate disease and regardless of vaccination status. Women of childbearing potential and men with female partners of childbearing potential were required to use a highly effective method of contraception for the duration of the treatment and for six weeks following the last dose. Exclusion criteria included pregnant or breastfeeding women, swallowing difficulties, known medical history of liver disease, receiving dialysis or known moderate to severe renal impairment (CKD stage 4 or 5), oxygen saturation <92% on room air (or their standard home oxygen supplementation), ALT>5 times upper limit of normal, known allergy to any study medication, or having received any other experimental agents within 30Ldays of first dose of study drug (use of contraindicated co-medications as defined in the Summary of Product Characteristics for molnupiravir^16^ and nirmatrelvir/ritonavir^17^ was not permitted; other medications were permitted at physician discretion following recommendations from the Liverpool COVID-19 Drug Interactions tool^18^). All participants provided written informed consent.

The study protocol was reviewed and approved by the UK Medicines and Healthcare Product Regulatory Agency (EudraCT 2020–001860-27) and West Midlands Edgbaston Research Ethics Committee (20/WM/0136).

### Molnupiravir, Nirmatrelvir/ritonavir and Standard of Care

Participants received molnupiravir (Lagevrio®) 800mg in combination with nirmatrelvir 300mg (Paxlovid®) boosted with ritonavir 100mg twice daily (12-hours apart +/- 4 hours, without regard to food) for 5 days as full doses at the start, with a de-escalation protocol reducing in decrements of molnupiravir to 600mg BD, then 400mg BD if required. Nirmatrelvir/ritonavir doses were fixed throughout the trial owing to the limited options for de-escalation. Participants were allowed flexibility in dosing by 4 hours; missed doses beyond this time were omitted. Standard of care included symptomatic relief such as antipyretics. No participant in the standard of care arm received any antiviral therapy. All participants attended for review on days 3, 5 and 11, bringing study medication bottles for drug accountability.

### Procedures

Participants with a lateral flow positive SARS-CoV-2 infection were screened against eligibility criteria, including presence and onset of symptoms within the previous 5 days. A total of 24 participants were randomly allocated, in a 2:1 ratio using block randomisation with no stratification, to either molnupiravir in combination with nirmatrelvir/ritonavir for 5 days or standard of care. Participants were recruited in cohorts of 6 (4 randomised to molnupiravir plus nirmatrelvir/ritonavir and 2 to standard of care), with review of safety and tolerability between cohorts. Participants were seen in clinic at baseline (day 1) and days 3, 5, 11 (+/- 2 days) and 29 (+/-2 days) with telephone follow up (and home collection of viral swabs) on days 2 and 4. A two-parameter Bayesian dose de-escalation model was utilised ([20] and supplement S1) to recommend the next dose level which targeted a safe dose with an additional DLT risk of 20% (the target interval of 15-25%) above standard of care. A dose was deemed unsafe if there was a ≥25% probability that the risk of toxicity is 30% higher than standard of care. As an additional stopping rule if the highest dose combination (800mg molnupiravir with nirmatrelvir/ritonavir) had a probability of greater than 47·5% of being unsafe (i.e. >47·5% probability that the risk of toxicity is 30% higher than standard of care), then the study would halt due to safety concerns. A Safety Review Committee reviewed each dose cohort of 6 participants, with recruitment paused only for the second review meeting (i.e. in the absence of any dose-limiting toxicities at this point, we would proceed to full recruitment of 24 participants as the probability of the combination being safe was very high, e.g. 95%). If dose-limiting toxicities (up to at least Day 11 of follow-up) were observed, de-escalation of the molnupiravir dose (to 600mg bd or 400mg bd) was considered by the Safety Review Committee, guided by safety data and the Bayesian dose-toxicity model. Although the Bayesian model could recommend de-escalation using DLTs, other adverse event (AE), vital signs, as well as any emerging data were considered by the Safety Review Committee.

### Outcomes

The primary outcome was dose-limiting toxicities (DLT) using CTCAE version 5 (grades 3 and above) up to and including day 11. Secondary outcomes for safety included AEs, SAEs, physical findings, vital signs (heart rate, blood pressure, respiratory rate, temperature & oxygen saturation), time to negative PCR and viral load reduction (estimated using the log_10_ mean pseudo-concentration of the 3 genes (N, ORF and S, if amplified)^20^), laboratory parameters including blood and urinary (with plasma, saliva, tears and nasal swabs for pharmacokinetics at day 1 and 5), ECG and the number of deaths and hospitalisations up to day 29.

#### Virological characterisation

Serial swabs (sampled from the oropharynx, then mid-turbinate space) were collected in DNA/RNA Shield (Zymo Research #R1100) as previously described^20^ at baseline (day 1), then days 2, 3, 4, 5 and 11 (with self-collection at days 2 and 4). Following extraction, viral RNA was quantified TaqPath COVID-19 RT-PCR Kit (ThermoFisher Scientific, Waltham, Massachusetts), with thresholds for each amplicon (S-gene, N-gene and OFR1) adjusted to give a threshold cycle of 32 with a control of 25 templates per reaction. Time of negativity within an amplicon was determined by the time of the first of two consecutive readings below the limit of detection (cycle threshold of 32 or more) where at least two amplicons were concordant. If all three amplicons differed, the median time to negative PCR was utilised. If only two amplicons were evaluable (e.g. if the third was censored), the later time of the two was utilised. Where only one amplicon was evaluable, time to negative PCR was censored at the last PCR measurement. In the event of S-gene amplification failure, the S-gene was considered censored at day 11 and the rules above applied. Viral titre was quantified by estimating a viral ‘pseudoconcentration’ (expressed as log10 copies of template per reaction) as previously described.^20^

#### Pharmacokinetic evaluation

Plasma was collected on Day 1 and Day 5 with sampling pre-dose (0h), 0·5, 1, 2 and 4 hours post-dose for quantification of ß-d-N4-hydroxycytidine (NHC; the active metabolite of molnupiravir), nirmatrelvir and ritonavir as previously described.^21,22^ Briefly, plasma extracts (obtained after protein precipitation using 3:1 (v/v) acetonitrile to plasma) were chromatographically separated using a reverse-phase Atlantis dC_18_ column (Waters UK) followed by LC-MS (AB Sciex 4500, Framingham, USA) analysis. The lower limits of quantification were as follows: NHC 2·5 ng/mL, nirmatrelvir 2·5 ng/mL and ritonavir 1·25 ng/mL. Non-compartmental analysis of truncated profiles on Day 1 and Day 5 were calculated using Phoenix WinNonlin v.8·3 software (Certara, USA) and presented as geometric means (95% CI).

We also measured concentrations of nirmatrelvir and ritonavir in non-plasma compartments on Day 5.^23^ Saliva was centrifuged from Salivette^TM^ swabs, chewed for 60 seconds. Nasal secretions were collected using Synthetic Absorptive Matrix (SAM) strips (Mucosal Diagnostics, UK) applied against the surface of the inferior turbinate of each nostril for 60 seconds – weights (to the nearest 0·1 mg) were recorded before and after sampling. Tears were collected using Schirmer Tear Test strips inserted in each lower eyelid for 5 minutes with approximate volumes (to the nearest µL) recorded using the graduated markings on the strip (1-35 µL). Drug extraction, and measurement using tandem LC-MS/MS has been previously described.^23^ The lower limit of quantitation for nirmatrelvir (saliva, tears, nasal secretions) was 2·5ng/mL, 0·04ng/sample, 0·04ng/sample), and for ritonavir in saliva this was 1·25ng/mL (ritonavir was not measured in tears or nasal secretions due to the very low levels seen in saliva, and because it is not an active component of the regimen for efficacy).

### Safety assessments

Safety was evaluated at specific time points throughout the trial using CTCAE version 5 with real time serious adverse event (SAE) reporting. Study visits at baseline and days 1, 3, 5 and 8 included collection of safety bloods (including full blood count, biochemistry, liver and renal function), clinical assessment and measurement of viral signs. At these visits, and additionally at days 2, 4, and 29, symptom, health and medication questionnaires were administered.

### Statistical analysis

We utilised an adaptive model-based dose-finding design based on^19^ with a plan for a total of up to 4 cohorts of 6 participants giving a maximum of 24 participants. All analyses were intention-to-treat apart from the safety analysis (which included only participants who received at least one dose of the allocated treatment). There was no imputation of missing data, data transformations (apart from viral load which used log_10_ viral load) or adjustment for multiplicity for any of the analyses. The primary and secondary analyses were conducted after all participants had been followed through day 29. The primary analysis of DLTs up to and including day 11 used a two parameter Bayesian dose de-escalation model with a prior estimate of the DLT risk for a participant on Standard of care assumed to be 10% with results presented as the number DLTs and posterior point estimates and 95% equal-tail credible intervals of the risk of a DLT for each dose (including Standard of care) (Supplementary data S1). As a secondary analysis the model was applied to all data available, i.e. DLTs up to day 29. Descriptive analyses of baseline characteristics and other endpoints were summarised using means, medians and proportions with corresponding IQRs or 95% CIs as appropriate. All analyses were reported according to CONSORT 2010 and the ICH E9 guidelines on Statistical Principles in Clinical Trials. All analyses were carried out in SAS v9·4 and Stata v16 except the Bayesian analyses, which were performed using packages available in R v4·0·2.

Viral load reductions from baseline were fitted to a Bayesian biexponential model ^24^ to represent “fast” decay at an initial stage of viral elimination and “persistent” decay at a second stage of viral elimination. The mixed model is used to account for variability in slopes between participants. In the primary pre-specified analysis model it was assumed that treatment only affects the fast decay (a subsequent sensitivity analysis also evaluated for effects on both phases of viral clearance). With t defined as time (days), the primary bi-exponential model is given by

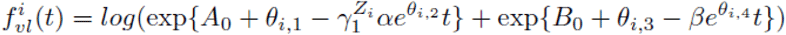

where A0 and α are the population intercept and slope respectively for the fast decay stage and B0 and β are the population intercept and slope respectively for the persistent decay stage. The parameter γ1 represents the treatment effect coefficient and Zi is the treatment indicator taking the value of 1 if patient I receives the experimental treatment and 0 otherwise. The vector θi = (θi,1, θi,2, θi,3, θi,4)T is a random effect specific to patient i. Full details of the model including the pre-specified prior distribution are given in Supplementary material S2. The posterior probability of the treatment effect coefficient being above 1 is used to declare whether there is a treatment effect on the viral slope. Based on the simulations, if this posterior probability is above 91% then the treatment effect can be declared controlling the rate of false positive conclusion at 10%.

## RESULTS

### Trial population

Between January and September 2023, 49 potential participants (Figure 1) were screened; 25 were excluded; 8 could not comply with clinic visits, 6 could not be re-contacted, 5 did not fulfil symptom criteria, 5 had negative lateral flow tests, and 1 could not meet the contraception requirements. The remaining 24 eligible participants were randomly assigned (2:1 to receive Combination antivirals or Standard of care) across 4 sequential cohorts of 6 participants each (giving a total of 16 randomised to combination antivirals and 8 to Standard of care), all of whom were included in the analysis.

**Figure 1:** CONSORT diagram

The baseline characteristics of participants were broadly similar with regard to age, sex, vaccination status, and body mass index (Table 1). Median interval from symptom onset was 4 days in the Combination arm, versus 3 days in the Standard of care arm. A total of 94% (15/16) of Combination participants completed the full treatment, with 1 starting treatment but withdrawing due to side effects (after receiving 4 doses out of 10 – i.e. after 2 days of full treatment). All 24 participants had their day 29 follow-up visit (+/- 2 days).

#### Primary analysis

No participants in any cohort experienced a DLT or a grade 3 or above adverse event (Table 2), consequently the Safety Review Committee confirmed all participants randomised to combination therapy should receive both drugs at full doses for 5 days. Data were analysed separately as well as pooled across cohorts to evaluate differences by arm. Bayesian model DLT point estimates, 95% credible interval and the target toxicity level of 20% over SoC are shown in Figure 2. After the 4 cohorts for data up to day 11, the 800mg molnupiravir dose in the Combination arm had an estimated DLT rate of 10·5% (equal-tail 95% credible interval of 3·8%–22·2%), with estimated 4·6% additional toxicity over Standard of care and a probability of additional toxicity 30% over SoC of 0%. Inclusion of DLTs recorded up to day 29 (supplementary data) into the model also supported a recommended Phase II dose of molnupiravir 800mg with nirmatrelvir (300mg)/ritonavir (100mg) BD for 5 days.

**Figure 2:** Bayesian estimates of dose-toxicity (primary endpoint) and virological efficacy Posterior estimates from participants in the combination (N = 16) and SoC (N=8) arms 2a Dose-Toxicity relationship (95% credible interval) to Day 11 (by intention-to-treat). 2b Viral titre (expressed as log10 ‘pseudocopies’/swab) derived from cycle threshold (CT) for three genes: N, S and ORF-1 as described in Methods. The mean CT of all three genes were used, with genes excluded if they did not amplify. 2c Biexponential model of virological clearance comparing slope parameter estimates of combination antiviral therapy versus Standard-of-care. The treatment coefficient (γ1) between both arms is 1.57 (95% confidence 1.22 – 2.07) for treatment effect, which is considered significant since it does not include 1. See supplementary data S2 and text.

#### Analysis of secondary endpoints

There were no deaths, hospitalisations or any new requirement for oxygen therapy for any participant. Nineteen out of the 24 participants in the trial reported at least one AE, with 79% reporting a worst grade of 1 (n=15) and 21% reported a worst grade of 2 (n=4). Fourteen of 16 (88%) on M+P and 5 of 8 (63%) SoC participants had at least one AE. Combination therapy was generally well-tolerated and Table 2 describes the frequencies of events across the groups by system organ class and CTCAE term. Although numbers were small, there did not appear to be any differences with the exception of dysgeusia (11/16; 69%), altered bitter taste (2/16; 12·5%) and tongue parasthesia (1/16; 6%) in participants receiving combination therapy compared with none in standard-of-care arm. No SAEs were reported.

The confirmed PCR negativity rate at both day 5 and 11 for combination therapy and Standard of care was 68·8% (n=11/16) and 62·5% (n=5/8) respectively with no significant differences observed at days 2, 3 and 4. Changes in viral titres offer greater precision for estimating treatment effect. Posterior estimates from a biexponential model of virological clearance over time are shown in Figure 2. The estimated mean of the treatment effect coefficient (γ1) is 1·57 (95% credible interval 1·22 – 2·07) for treatment effect. The posterior probability of the treatment effect coefficient being positive is >99·9%. This is above the pre-specified threshold of 91%, i.e. the treatment effect on the fast decay slope can be concluded. The sensitivity analysis of the model with the treatment effect on both fast and delay decay suggest a high confidence (87%) that the treatment effect coefficient for the fast decay is above 1.

### Pharmacokinetics

#### Plasma pharmacokinetics

For NHC, a total of 109 samples were obtained from 11 participants at Days 1 and 5 (10 evaluable, 1 subject with missing samples on Day 5 was excluded). Geometric mean (95% CI) AUC_0-4_ was 7054 (5581-8917) ng·h/mL on Day 1, and 7932 (6565-9585) ng·h/mL on Day 5. Geometric mean (95% CI) Cmax was 3118 (2435-3992) ng/mL on Day 1, and 3335 (2764-4025) ng/mL on Day 5. T_max_ was 2·00 (range 1·00-4·00) hours and there was no evidence of any significant accumulation between Days 1 and 5.

For nirmatrelvir and ritonavir, a total of 153 samples were obtained from 16 participants at Days 1 and 5. For nirmatrelvir, geometric mean (95% CI) AUC_0-4_ was 13,998 (10,671-18,361) ng·h/mL on Day 1, and 20,884 (17,624-24,747) ng·h/mL on Day 5. Geometric mean (95% CI) Cmax was 5430 (4352-6775) ng/mL on Day 1, and 6414 (5412-7602) ng/mL on Day 5. T_max_ was 2·00 (range 2·00-4·00) hours. For ritonavir, geometric mean (95% CI) AUC_0-4_ was 1076 (645-1975) ng·h/mL on Day 1, and 3063 (2327-4032) ng·h/mL on Day 5. Geometric mean (95% CI) Cmax was 438 (295-651) ng/mL on Day 1, and 1129 (852-1498) ng/mL on Day 5. T_max_ was 2·00 (range 2·00-4·00) hours. There was an approximately 50% increase in nirmatrelvir, and an approximately three-fold increase in ritonavir exposure (P=0·001 for both; Wilcoxon test) at Day 5 compared with Day 1

#### Non-plasma pharmacokinetics (Figure 3)

A total of 74 saliva, 73 tear and 74 nasal swabs were analysed from 15 participants. All saliva samples yielded measurable concentrations of nirmatrelvir, with geometric mean (95% CI) AUC_0-4_ and Cmax of 3885 (2839-5317)ng·h/mL and 1294 (952-1758)ng/mL. Nirmatrelvir was detected in all tear samples, with geometric mean (95% CI) AUC_0-4_ and Cmax of 17,558 (12,769-24,143)ng·h/mL and 6789 (4892-9421)ng/mL. Nirmatrelvir was also detected in all nasal secretions, with geometric mean (95% CI) AUC_0-4_ and Cmax of 13,150 (9,060-19,086)ng·h/mL and 4713 (3198-6946)ng/mL. For ritonavir, 1 of 15 pre-dose saliva samples at Day 5 was below the lower limit of quantitation. The geometric mean (95% CI) AUC_0-4_ and Cmax were 28 (20-39)ng·h/mL and 10 (7-16)ng/mL. Given that ritonavir is not an active component of the regimen for efficacy, and that concentrations in saliva were low, we did not measure ritonavir in tears or nasal secretions. Compartmental penetration of nirmatrelvir calculated using plasma:compartment geometric mean AUC_0-4_ ratios (95%CI) was 0·18 (0·14-0·24) for saliva, 0·91 (0·69-1·19) for tears and 0·65 (0·50-0·83) for nasal secretions. The corresponding ratio for ritonavir was 0·009 (0·006, 0·013).

**Figure 3:** Pharmacokinetics of NHC, Nirmatrelvir and Ritonavir

## DISCUSSION

Better therapeutic options are an essential pillar of the 100 Day Mission.^25^ In AGILE CST-8 we have shown that the combination of oral molnupiravir plus nirmatrelvir/ritonavir for 5 days is safe and reasonably well-tolerated at full doses of both drugs. We did observe transient taste, and mild gastrointestinal disturbance with combination therapy, which are well-recognised dose dependent effects with ritonavir and it is worth noting that doses of ritonavir when administered with nirmatrelvir are double those usually given for boosting HIV treatment.

Importantly, plasma concentrations of NHC were generally within the range reported for our previous AGILE CST-2 trial.^26^ There was evidence of accumulation of nirmatrelvir, with plasma AUC approximately 50% higher at Day 5 compared with Day 1, and geometric mean Cmax concentration on Day 1 was 5430ng/mL in this study (compared with 2210 ng/mL reported in the drug label following a single dose of nirmatrelvir (300mg) plus ritonavir (100mg) to healthy volunteers^17^). To evaluate potential impact on sterilisation we have previously reported saliva, nasal, and tear NHC concentrations of 3%, 21%, and 22% that of plasma.^26^ In this study we observed saliva, nasal, and tear nirmatrelvir concentrations of 18%, 65%, and 91% that of plasma, despite low penetration of ritonavir into saliva.

Although our use of model-based estimates of risk for dose-limiting toxicity was able to accelerate decision making, the limited numbers of participants makes continued evaluation in Phase IIb necessary (this is a pre-requisite for any follow-on study regardless of statistical approach). Many questions surround the value of viral load measurements, given the lack of correlation with symptoms^27–29^ including rebound,^30^ and an unclear correlation with burden of infection in the lower respiratory tract. Despite our small sample size, the use of model-based estimation of viral clearance was able to detect a significant difference in favour of the treatment arm.

It is important to adequately characterise the safety of combination antivirals for SARS-CoV-2 infection, not only to allow testing in severe or persistent infections with currently circulating variants, but also have ready-to-test regimens available for rapid evaluation in the event of any new variant, or new zoonotic transmissions where existing therapies become compromised. The approach taken here is pragmatic, and cost-effective and could be extended to other antiviral combinations, including for emerging outbreaks beyond SARS-CoV-2.

## Contributors

SHK, GG, RF, TF, TJ, PM, LW contributed to study design. SHK, GG, GS, JN contributed to data analysis and interpretation. RF, SA, CJE led clinical conduct as principal investigators of the clinical sites. RL, RF, LW, CJE, SA, AB participated in clinical assessment and data collection. LJE, LD, VS, WG, CH, KB, AA contributed to study bioanalysis. EK, HER, MT, CM, JD, JG, JC contributed to study management and execution. DGL, AO, MJ contributed to the design of the AGILE platform. The manuscript was written by the authors, with SHK and GG as the overall lead authors. No one who is not an author contributed to writing the manuscript. All authors had full access to the data and GS and JN directly accessed and verified the underlying data reported in the manuscript. The authors assume responsibility for the accuracy and completeness of the data and for the fidelity of the trial to the protocol.

## Data Sharing Statement

The AGILE Trial Steering Committee will consider all reasonable requests by health-care providers, investigators, and researchers to provide anonymised data to address specific scientific or clinical objectives. The AGILE investigators are committed to reviewing requests from researchers for access to clinical trial protocols, de-identified patient-level clinical trial data, and study-level clinical trial data. Data will be assigned a DOI through deposition in the University of Liverpool Research Data Catalogue and shared under a Data Transfer agreement (or equivalent e.g. as part of a research collaboration agreement or confidentiality disclosure agreement).

## Acknowledgements

**<u>AGILE CST-8 Study Group</u>**

<u>AGILE Independent Trial Steering Committee</u>

Nicholas Paton (chair), Fred Hayden, Janet Darbyshire, Amy Lucas, Ulrika Lorch.

<u>AGILE independent Data Monitoring and Ethics Committee</u>

Andrew Freedman (chair), Richard Knight, Steven Julious

<u>Liverpool School of Tropical Medicine</u>

Thomas Edwards, Christopher Myerscough

<u>Southampton Clinical Trials Unit</u>

Nuala Tainton, Oliver Edwards, Kerensa Thorne

<u>University of Liverpool</u>

Sujan Dily Penchala, Rachel Carter

<u>NIHR Liverpool Clinical Research Facility</u>

Callum Kelly, Kate Dodd, Callum Docherty

<u>NIHR Manchester Clinical Research Facility</u>

Charlotte Boe

<u>NIHR Southampton Clinical Research Facility</u>

Alasdair Munro, Dan Owens, Mihaela Pacurar, Gavin Babbage, Liza Shiner-Clark

We are deeply grateful to all participants for taking part in this trial. We are also grateful to Debbie Ellis, Emma Tilt (Southampton Clinical Trials Unit), and Laura Bradley, Karen Jennings-Wilding (University of Liverpool), and May Lwin, Thomas Reed, Tom Durham, Simone Paulson, Giorgio Bartalucci, Mihai-Adrian Panainte, Mayu Otuska, Lara Marie Narvios, Juliza Ann Vidal, Cristina Zilio, Natasha Tantony, Satwinder Virdee, Ifeoma Ezewudo, Patricia Acosta Carrazco, Noshin Daula, Alice Sebastian, Caroline Grabau, Danny Pratt, Seena Joseph, Angelica Salgado, Pheobe Wamban, Xy-Za Albana, Emma Greene (Southampton Clinical Research Facility).

