## Supplementary information for "A randomised-controlled Phase I de-escalation trial of Molnupiravir and Nirmatrelvir/Ritonavir combination for mild-moderate SARS-CoV-2 infection" (from another version of this preprint: version 1, 2025-05-02)

### Supplementary Data

**Table of Contents**

S1 Bayesian Model Page 2

S2 Analysis of Viral Clearance using a Biexponential model Page 6

**S1 Bayesian Model**

### Bayesian Dose De-escalation Model for CST-8

The Bayesian combination-toxicity model is given by

$$\psi(\tilde{d}_j, \theta_1, \theta_2) = \frac{\exp(\theta_1 + \theta_2 \tilde{d}_j)}{1 + \exp(\theta_1 + \theta_2 \tilde{d}_j)} \quad (1)$$

where $\tilde{d}_j, j = 0, 1, 2, 3$ are the standardized levels of combinations obtained through prior estimates of the DLE probabilities at the respective combinations $\hat{p}_j^{(0)}$, and $\theta_1, \theta_2$ are the model parameters with a prior distribution.

The standardized levels of combinations are obtained as

$$\tilde{d}_j = \frac{\text{logit}(\hat{p}_j^{(0)}) - \hat{\theta}_1^{(0)}}{\hat{\theta}_2^{(0)}}$$

where $\hat{\theta}_1^{(0)}$, $\hat{\theta}_2^{(0)}$ are prior point estimates of the model parameters, with $\tilde{d}_0 = 0$.

The study proceeds in the cohort of 6 patients (4 experimental : 2 control) with the starting combination being the highest one ($j = 3$). After each cohort of patients and their DLTs outcomes being evaluated, the model is updated and the posterior distributions of the model parameters are used to derive the estimates of the toxicity risk at each combination. Given the posterior distribution, the design uses the following rules for the decision-making:

- The combination $j$ is unsafe and is not recommended for the next cohort of patients if

$$\mathbb{P}\left(p_j - p_0 > 0.30\right) > 25\%$$

- If all combinations are found to be unsafe, then the study is stopped earlier for safety.
- The highest combination is deemed unsafe and the study is recommended to be halted due to safety concerns if

$$\mathbb{P}\left(p_3 - p_0 > 0.30\right) > 47.5\%$$

For the setting with 3 experimental combinations, the prior parameters were calibrated using the scenario 1 and scenario 2 given in Table 1 over a grid of values of each of the hyperparameters. The following parameters were found to result in good operating characteristics over all two scenarios: $(\theta_1, \log(\theta_2)) \sim \mathcal{N}(\mu, \Sigma)$ where $(\mu_1, \mu_2) = (\text{logit}(0.1), 0.00)^{\text{T}}$ is the vector of means and

$$\Sigma = \begin{bmatrix} 0.30 & 0 \\ 0 & 0.75 \end{bmatrix}.$$

using the spacing between prior probabilities of 0.10, the vector of standardized dose levels of $\tilde{d} = (0.000, 0.557, 0.927, 1.231)$.

The proportion of each dose selection under 3 scenarios with $N = 24$ are given in Table 1.

Table 1: Proportion of each dose selection under the calibrated model for $N = 24$. The correct selections are in bold. Results are based on 2000 simulations.

| | $d_0$ | $d_1$ | $d_2$ | $d_3$ | Terminations for Safety | Av SS |
|---|---|---|---|---|---|---|
| | Scenario 1 | | | | | |
| Toxicity | 10% | 30% | 40% | 50% | | |
| Selection | – | 0.01 | 0.15 | 0.12 | **0.72** | 14.6 |
| | Scenario 2 | | | | | |
| Toxicity | 10% | 12% | 15% | 30% | | |
| Selection | – | 0.00 | 0.14 | **0.70** | 0.16 | 19.8 |
| | Scenario 3 | | | | | |
| Toxicity | 10% | 12% | 15% | 20% | | |
| Selection | – | 0.00 | 0.06 | **0.89** | 0.05 | 19.3 |

#### Escalation Strategies

Illustration of the escalation decision after the first cohort of patients (2 patients on the control, 4 patients on the highest combination) if 0 DLTs out of 2 patients, and X DLTs out of 4 patients (for varying X). For each case, a table with the following quantities is presented

- Estimated Toxicity (Mean)
- Overdose Probability = $\mathbb{P}(p_j - p_0 \geq 30\%)$
- Target Probability = $\mathbb{P}(p_j - p_0 \in (15\%, 25\%))$
- Safe Probability = $\mathbb{P}(p_j - p_0 \leq 25\%)$

Note that the Probabilities are formulated in terms of the *excessive toxicity* (over the control).

##### 0 DLTs

| | SoC | $d_1$ | $d_2$ | $d_3$ |
|---|---|---|---|---|
| Mean Toxicity | 8.50 | 11.94 | 15.09 | 18.22 |
| Overdose | 0.00 | 0.00 | 0.57 | 3.87 |
| Target | 0.00 | 0.31 | 7.20 | 12.84 |
| Safe | 0.00 | 100.00 | 98.53 | 93.42 |

Next Recommended Combination: Combination $d_3$

##### 1 DLTs

| | SoC | $d_1$ | $d_2$ | $d_3$ |
|---|---|---|---|---|
| Mean Toxicity | 9.99 | 15.55 | 20.88 | 26.09 |
| Overdose | 0.00 | 0.00 | 3.83 | 13.53 |
| Target | 0.00 | 3.31 | 16.88 | 22.71 |
| Safe | 0.00 | 99.92 | 92.23 | 80.27 |

Next Recommended Combination: Combination $d_3$

##### 2 DLTs

| | SoC | $d_1$ | $d_2$ | $d_3$ |
|---|---|---|---|---|
| Mean Toxicity | 11.54 | 21.12 | 30.54 | 39.12 |
| Overdose | 0.00 | 0.84 | 17.55 | 38.66 |
| Target | 0.00 | 13.17 | 27.94 | – |
| Safe | 0.00 | 97.40 | 73.49 | 51.56 |

Next Recommended Combination: Combination $d_2$ (De-escalate)

#### 3 DLTs

| | SoC | $d_1$ | $d_2$ | $d_3$ |
|---|---|---|---|---|
| Mean Toxicity | 12.28 | 29.48 | 45.81 | 58.24 |
| Overdose | 0.00 | 9.96 | 53.95 | 73.46 |
| Target | 0.00 | 31.63 | – | – |
| Safe | 0.00 | 80.61 | 37.46 | 20.41 |

Next Recommended Decision: Stop trial earlier for safety due to the highest combination having a high risk of being unsafe.

#### S2 Analysis of Viral Clearance using a Biexponential model

**Analysis of CST-8 data using bi-exponential model**

##### Purpose

This document provides the statistical analysis of the CST-8 trial evaluating preliminary evidence of virological efficacy in patients receiving the experimental treatment, Molnupiravir 800mgTwice a day (BD) in combination with Paxlovid® (300mg nirmatrelvir + ritonavir 100mg) twice a day(BD) for 5 days, compared with patients receiving the standard of care, Paxlovid® (300mg nirmatrelvir + ritonavir 100mg) twice a day (BD) for 5 days. The trial recruited 16 patients to the experimental treatment arm and 8 patients to the standard of care treatment arm.

##### Endpoint

The endpoint of interest is the difference in slopes of viral load between treatment groups. To calculate viral load, the following components of cycle threshold (CT) were collected for three genes Gene-N, Gene-S and ORF1. Estimates of the viral load at each time point were found by taking the mean of the 3 genes viral load pseudo-concentration values assuming they have all 'amplified'. If one gene has not amplified then viral load was calculated as the mean of the other two genes. A log10 transform was then applied to give the final viral load measurement at a given timepoint.

##### Descriptive analysis

Table 1 shows the baseline characteristics for the 24 patients recruited to the trial. Randomisation was not stratified by patient characteristics and we see that allocation is reasonably split across participants.

| Characteristic | Paxlovid alone (n=8) | Molnupiravir + Paxlovid (n=16) |
|---|---|---|
| **Sex** | | |
| Male | 1 | 6 |
| Female | 7 | 10 |
| **Age** mean (sd) | 37.5 (12.7) | 42.3 (16.7) |
| **Vaccination status** | | |
| Vaccinated against COVID-19 | 6 | 14 |
| Not vaccinated | 2 | 2 |
| **Day of symptoms onset** mean (sd) | 3.3 (1.39) | 3.8 (0.75) |

Table 1: Table of baseline characteristics of patients upon entering the trial, split by treatment arm.

Table 2 shows the mean and standard deviation of the viral load endpoint, split by treatment arm and day of test. In this case, day 1 is the baseline measurement, before first treatment is administered and as per protocol, day 11 tests have a 2 day window of acceptability. We also provide the change in baseline of this endpoint and the number of missing observations for each test day. Further, Figure 1 shows a boxplot of the viral load measurements and Figure 2 shows a boxplot of the change from baseline in viral load measurements. It appears that the Molnupiravir + Paxlovid treatment group has a lower mean log viral load than for the Paxlovid alone treatment arm.

##### Primary analysis

A bi-exponential model has been fitted to the log10 viral load measurements to represent "fast" decayat an initial stage of viral elimination and "persistent" decay at a second stage of viral elimination under

| Measurement | Day 1 (baseline) | 2 | 3 | 4 | 5 | 11 (±2 days) |
|---|---|---|---|---|---|---|
| **log10 viral load, mean (sd)** | | | | | | |
| Paxlovid alone | 6.07(2.91) | 3.60(2.39) | 1.85(2.39) | 1.30(1.76) | 1.03(1.98) | 0.60(1.98) |
| Molnupiravir + Paxlovid | 5.08(1.81) | 3.04(2.47) | 1.84(2.08) | 0.84(1.96) | 0.10(1.03) | -0.48(1.03) |
| **log10 viral load change from baseline, mean (sd)** | | | | | | |
| Paxlovid alone | 0.00(0.00) | -2.47(2.01) | -4.23(2.40) | -4.77(2.59) | -5.04(2.80) | -5.47(2.80) |
| Molnupiravir + Paxlovid | 0.00(0.00) | -1.95(1.59) | -3.23(2.32) | -4.24(2.28) | -4.97(1.72) | -5.55(1.72) |
| **Number of missing observations** | | | | | | |
| Paxlovid alone | 0 | 0 | 0 | 0 | 0 | 0 |
| Molnupiravir + Paxlovid | 0 | 1 | 0 | 0 | 0 | 0 |

Table 2: Mean and standard deviation of log10 viral load raw values, change from baseline and number of missing observations at day 1,2,3,4,5 and day 11 of treatment split by treatment arm.

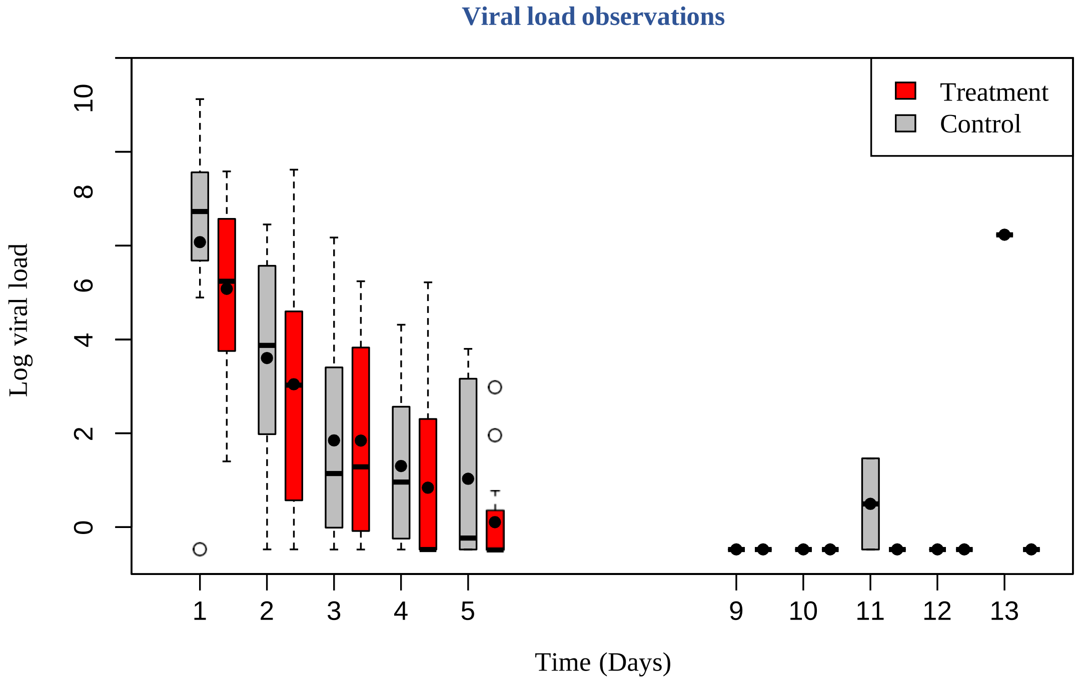


Figure 1: Box plot of viral load measurement split by day and treatment group. Solid points give means as in Table 2 and lines show medians.

low densities. A mixed model is used to account for variability in viral load slopes between patients. It is assumed that treatment only affects the fast decay, which is our primary analysis model (1) (but it could be that persistent decay is also affected by treatment and this will be considered for sensitivity in model (3)). With $t$ defined as time (days), the bi-exponential model is given by

$$f_i^{v}(t) = log(\exp\{A_0 + \theta_{i,1} - \gamma_1^{Z_i} \alpha e^{\theta_{i,2}} t\} + \exp\{B_0 + \theta_{i,3} - \beta e^{\theta_{i,4}} t\}) \quad (1)$$

where $A_0$ and $\alpha$ are the population intercept and slope respectively for the fast decay stage and $B_0$ and $\beta$ are the population intercept and slope respectively for the persistent decay stage. The parameter $\gamma_1$ represents the treatment coefficient and $Z_i$ is the treatment indicator taking the value of 1 if patient $i$ receives the experimental treatment and 0 otherwise. The vector $\boldsymbol{\theta}_i = (\theta_{i,1}, \theta_{i,2}, \theta_{i,3}, \theta_{i,4})^T$ is a random

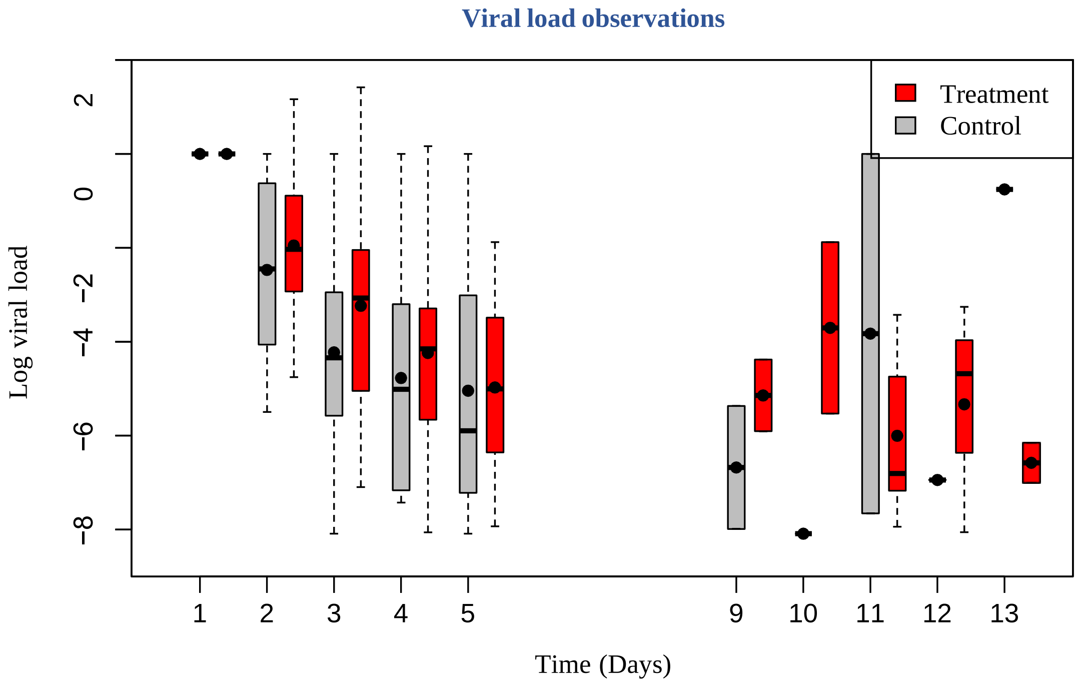


Figure 2: Box plot of change from baseline in viral load measurement split by day and treatment group. Solid points give means as in Table 2 and lines show medians.

effect specific to patient $i$.

Missing values of viral load measurements were handled by simply removing these observations from the analysis and were not imputed. The mixed models framework can be used in this setting becausethe bi-exponential trajectory in model (1) can be fitted to any patient with four or more log10 viral load measurements. We planned that patients with three or less viral load measurements would be left out of the analysis, however this was not the case for any patient. Viral load measurements below the limit of detection (1/3 RNA copies per ml) are considered censored. Let $\delta_i(t)$ be the indicator function taking the value of 1 at time $t$ if the viral load measurement for patient $i$ is censored and 0 otherwise. The following hierarchical model was fitted:

$$P(\delta_i(t)) = \begin{cases} 1 & \text{if } log(VL_i(t)) \leq log_{10}(1/3) \\ 0 & \text{otherwise} \end{cases}$$
$$log(VL_i(t)) \sim N(f^i(t), \sigma^2)$$
$$\boldsymbol{\theta}_i \sim (\mathbf{0}, \Sigma)$$

The prior distributions were initially chosen to be as in Watson et al. [2022]. We observed that uncertainty for most parameters was high. This was particularly true for parameters involved in the slopes e.g $\alpha$ and $\beta$. With Watson et al. [2022] as a starting point, we adjusted the uncertainty around some parameters to be smaller to avoid difficulty fitting the model to the data under smaller sample sizes. The following priors for the primary analysis were therefore used:

$$A_0 \sim N(5, 0.5), \alpha \sim N(0.979, 0.1), \gamma_1 \sim N(1, 0.2),$$
$$B_0 \sim N(-0.4, 0.5), \beta \sim N(-0.003, 0.1), \sigma^2 \sim Exp(1), \Sigma \sim W_5(I_4). \quad (2)$$

where $W_5(I_4)$ is a Whishart distribution with 5 degrees of freedom and $I_4$ representing the $4 \times 4$ identity matrix. Figure 3 shows the prior and posterior time curves along with 95% confidence intervals when the CST-8 data is fitted to model (1).

The output of the bi-exponential model is given in Table 3. The main analysis focuses on theparameter $\gamma_1$ which will be equal to 1 if there is no difference in slopes of viral load between treatment

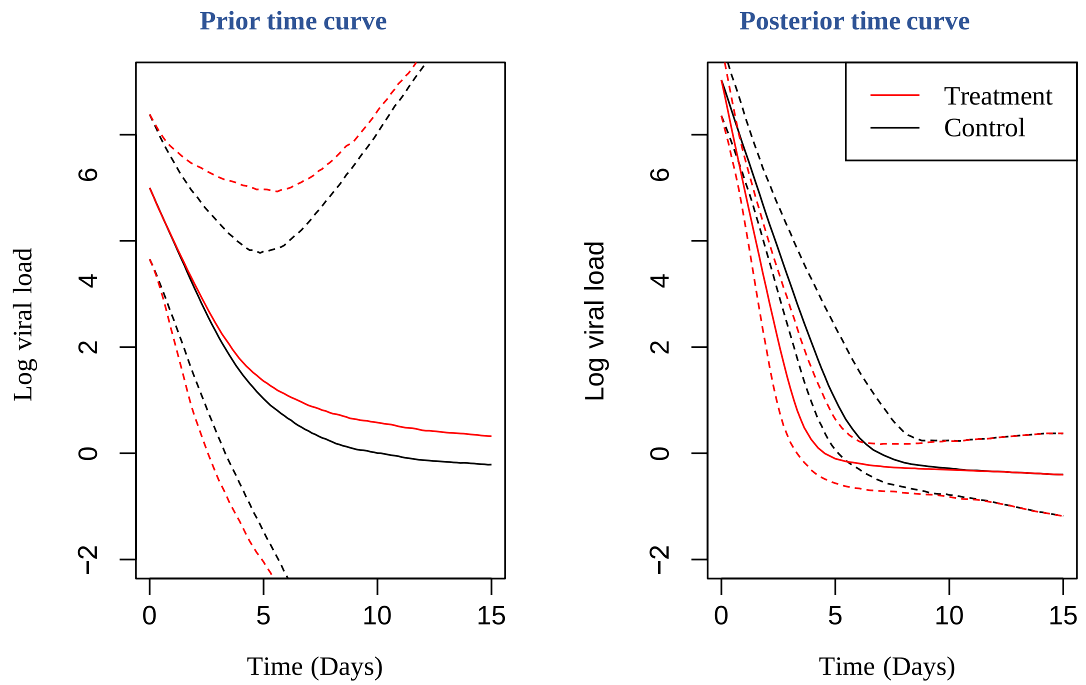


Figure 3: Prior and posterior time curves for model (1). The median curves for treatment and control lines are given by solid lines and the dashed lines represent the 95% credible intervals.

arms. The posterior samples for $\gamma_1$ have mean 1.57 the 95% confidence (1.22, 2.07) which does not contain 1, so there is good evidence that experimental treatment is working well. Log viral load difference between treatment arms at day 5, given by $f_{vl}(5; Z_i = 0) - f_{vl}(5; Z_i = 1)$, will be 0 if treatments have similar viral load slopes. Posterior estimates have mean 1.16 and 95% confidence interval (0.32, 2.25) which does not contain 0. Again, this provides good evidence that the experimental treatment is working better than control. Posterior samples of these treatment effects are plotted in Figure 6.

| **Parameter** | **Mean** | **95% confidence interval** |
|---|---|---|
| $\gamma_1$ | 1.59 | (1.22, 2.07) |
| $P(\gamma_1 > 1)$ | 1.00 | - |
| $A_0$ | 7.02 | (6.35, 7.73) |
| $\alpha$ | 1.27 | (0.86, 1.66) |
| $B_0$ | -0.13 | (-0.74, 0.46) |
| $\beta$ | 7.02 | (6.35, 7.73) |
| $\sigma^2$ | 1.49 | (1.13, 2.02) |
| $f_{vl}(5; Z_i = 0) - f_{vl}(5; Z_i = 1)$ | 1.16 | (0.32, 2.25) |
| $P(f_{vl}(5; Z_i = 0) > f_{vl}(5; Z_i = 1))$ | 1.00 | - |

Table 3: Output for the primary bi-exponential model with a single treatment parameter.

Table 4 shows the posterior probability thresholds which are needed to control error rates for each ofthe endpoints considered. For the first analysis based on the parameter $\gamma_1$, given the output in Table 3, this suggests that at a significance level of $\alpha = 0.1$, we reject the null hypothesis of $H_0 : \gamma_1 = 1$ in favour of the alternative $H_A : \gamma_1 > 1$. Based on the endpoint considering the difference in log viral load between treatment arms at day 5, for significance level $\alpha = 0.1$ we reject the null hypothesis $H_0 : f_{vl}(5; Z_i = 0) = f_{vl}(5; Z_i = 1)$ in favour of the alternative $H_A : f_{vl}(5; Z_i = 0) > f_{vl}(5; Z_i = 1)$.

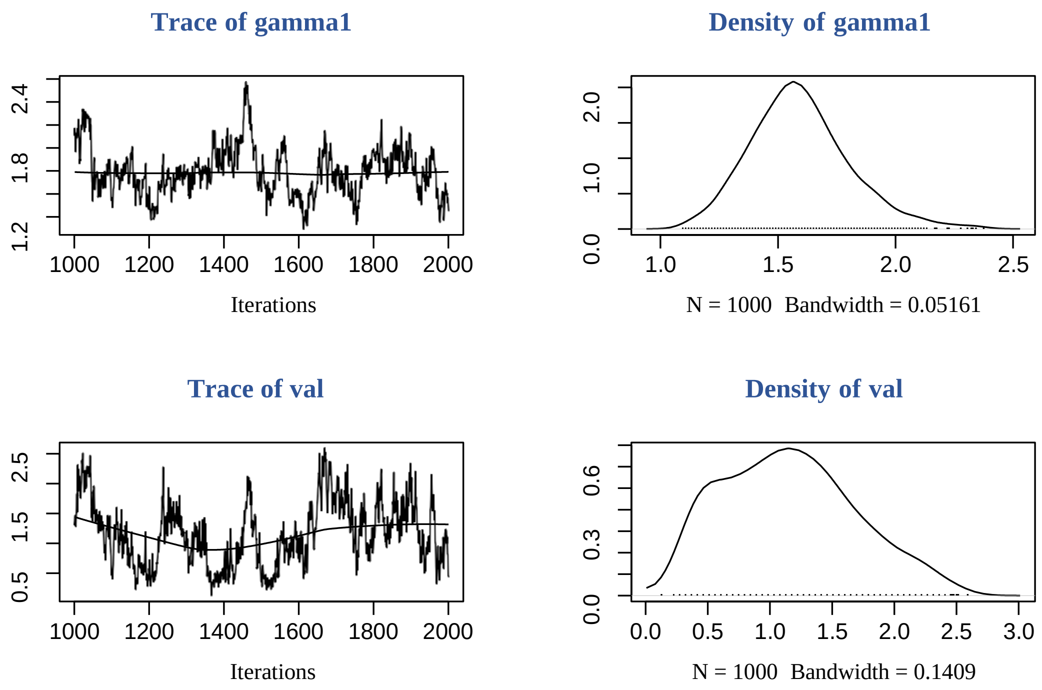


Figure 4: Posterior distributions of the treatment effects $\gamma_1$ and $f_{vl}(5; Z_i = 0) - f_{vl}(5; Z_i = 1)$ when fitting model (1) to the data.

| **error rate** | **P($\gamma_1 > 1$)** | **P($f_{vl}(5; Z_i = 0) > f_{vl}(5; Z_i = 1)$)** |
|---|---|---|
| 0.1 | 0.91 | 0.91 |
| 0.2 | 0.81 | 0.81 |
| 0.3 | 0.72 | 0.72 |

Table 4: Posterior probability thresholds for controlling type 1 error rates under the one treatment parameter (primary) model.

#### Sensitivity analysis

We also considered a two-treatment parameter bi-exponential model where treatment affects both fast decay and persistent decay slopes. The two treatment parameter model is given by:

$$f^i_{vl}(t) = log(\exp\{A_0 + \theta_{i,1} - \gamma_1^{z_i} \alpha e^{\theta_{i,2}} t\} + \exp\{B_0 + \theta_{i,3} - \gamma_2^{z_i} \beta e^{\theta_{i,4}} t\}). \quad (3)$$

In comparison with model (1), the difference here is the inclusion of the parameter $\gamma_2$. The same hierarchical model structure as for the first model will be used and the follwoing prior distribution isused

$$A_0 \sim N(5, 0.5), \alpha \sim N(0.979, 0.1), \gamma_1 \sim N(1, 0.2), \gamma_2 \sim N(1, 0.2),$$
$$B_0 \sim N(-0.4, 0.5), \beta \sim N(-0.003, 0.1), \sigma^2 \sim Exp(1), \Sigma \sim W_5(I_4). \quad (4)$$

Note that this prior is the same as (2) for the primary analysis but with the inclusion of the prior for $\gamma_2$.

The output from model 2 is shown in Table 5 and the prior and posterior time curves for this modelare shown in Figure 5. A positive value of $\gamma_2$ indicates that the log viral load measurements are greaterin the control group than for the treatment group during the persistent decay phase. However, the mean estimate for the parameter $\gamma_2$ is 0.34 with 95% confidence interval (−0.21, 1.35). The confidence interval includes 1 which is not significant and indicates that this parameter is not needed and that model (1) is sufficient. Posterior samples of the treatment effects are plotted in Figure 5.

| Parameter | Mean | 95% confidence interval |
|---|---|---|
| $\gamma_1$ | 1.41 | (0.70, 2.09) |
| $P(\gamma_1 > 1)$ | 0.87 | - |
| $\gamma_2$ | 0.34 | (-0.21, 1.35) |
| $P(\gamma_2 > 1)$ | 0.07 | - |
| $A_0$ | 6.93 | (6.12, 7.93) |
| $\alpha$ | 1.42 | (0.86, 1.87) |
| $B_0$ | -0.56 | (-1.19, -0.01) |
| $\beta$ | 6.93 | (6.12, 7.93) |
| $\sigma^2$ | 1.41 | (1.04, 1.97) |
| $f_{vl}(5; Z_i = 0) - f_{vl}(5; Z_i = 1)$ | 0.95 | (-1.20, 2.40) |
| $P(f_{vl}(5; Z_i = 0) > f_{vl}(5; Z_i = 1))$ | 0.90 | - |

Table 5: Output for the sensitivity analyses fitting data to a bi-exponential model with two treatment effect parameters

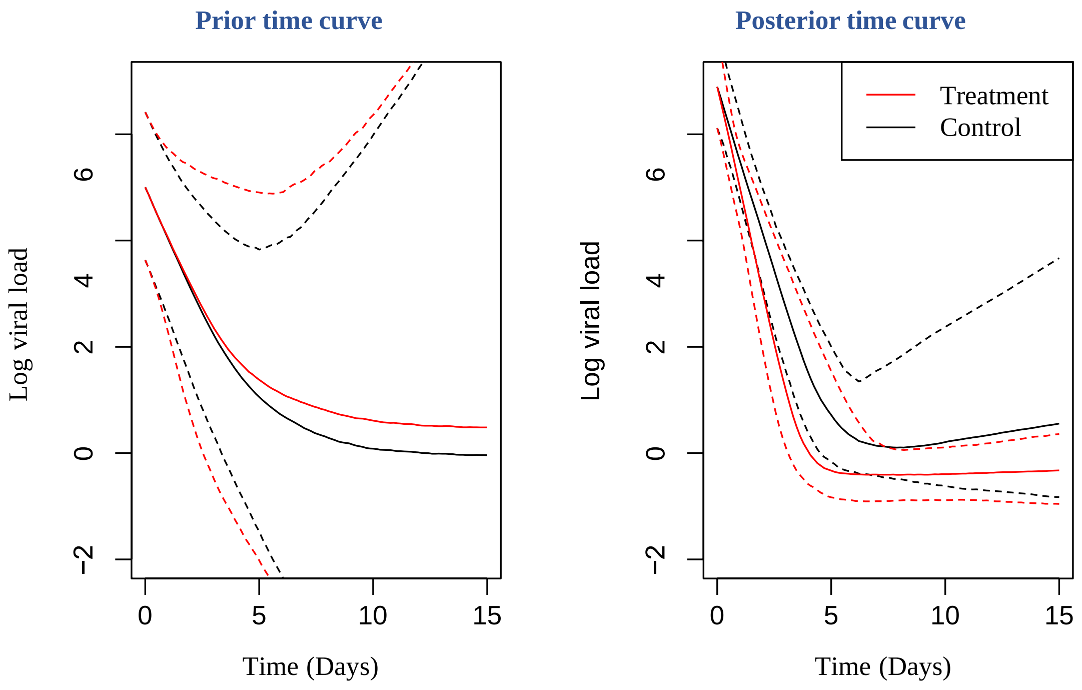


Figure 5: Prior and posterior time curves for model (3). The median curves for treatment and control lines are given by solid lines and the dashed lines represent the 95% credible intervals.

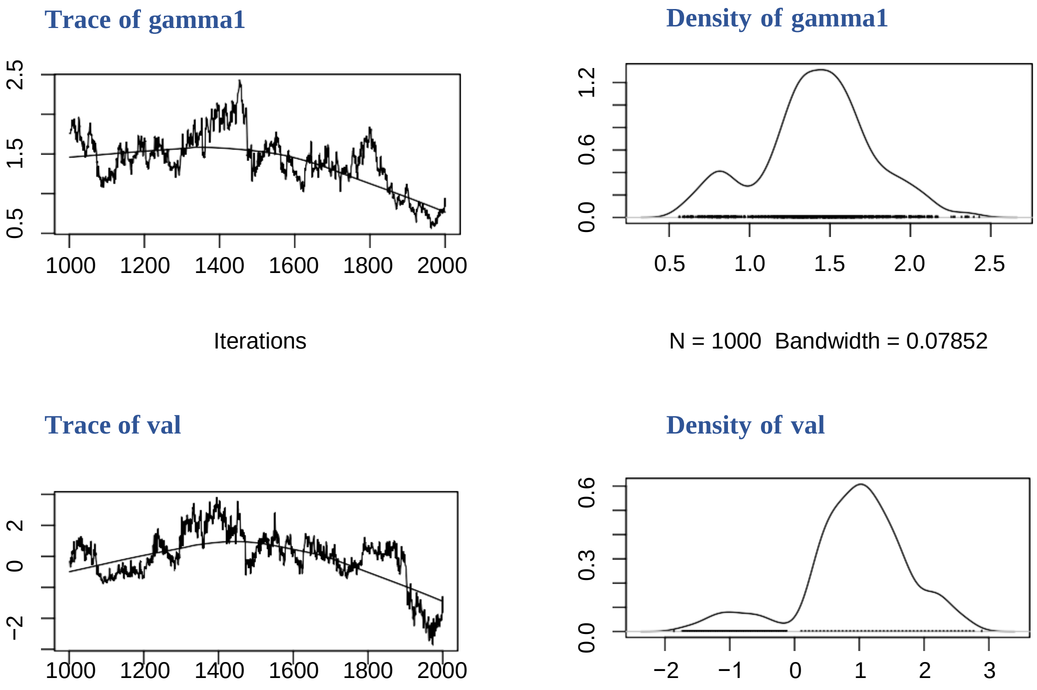


Iterations N = 1000 Bandwidth = 0.1696

Figure 6: Posterior distributions of the treatment effects $\gamma_1$ and $f_{yl}(5; Z_i = 0) - f_{yl}(5; Z_i = 1)$ when fitting model (3) to the data.
